# Genetic overlap between idiopathic pulmonary fibrosis and COVID−19

**DOI:** 10.1101/2021.12.08.21267459

**Authors:** Richard J Allen, Beatriz Guillen-Guio, Emma Croot, Luke M Kraven, Samuel Moss, Iain Stewart, R Gisli Jenkins, Louise V Wain

## Abstract

Genome-wide association studies (GWAS) of coronavirus disease 2019 (COVID-19) and idiopathic pulmonary fibrosis (IPF) have identified genetic loci associated with both traits, suggesting possible shared biological mechanisms. Using updated GWAS of COVID-19 and IPF, we evaluated the genetic overlap between these two diseases and identified four genetic loci (including one novel) with likely shared causal variants between severe COVID-19 and IPF. Although there was a positive genetic correlation between COVID-19 and IPF, two of these four shared genetic loci had an opposite direction of effect. IPF-associated genetic variants related to telomere dysfunction and spindle assembly showed no association with COVID-19 phenotypes. Together, these results suggest there are both shared and distinct biological processes driving IPF and severe COVID-19 phenotypes.

## Introduction

Coronavirus disease 2019 (COVID-19) is an infectious disease potentially leading to long lasting respiratory symptoms and has resulted in over 4 million deaths worldwide. Idiopathic pulmonary fibrosis (IPF) is a chronic interstitial lung disease (ILD) characterised by an aberrant response to alveolar injury leading to progressive scarring of the lungs. Individuals with ILD are at a higher risk of death from COVID-19^1^.

Large genome-wide association studies (GWAS) have identified multiple genetic signals associated with severe COVID-19^2^, including a signal within the *DPP9* gene that is also associated with increased IPF risk^3^. GWAS have identified 20 genome-wide significant signals of association with IPF risk^4,5^ with the largest genetic risk factor being a common variant located in the promoter region of *MUC5B* (rs35705950, odds ratio>4). Previous analyses suggest IPF is a causal risk factor for severe COVID-19 but noted that the effect of rs35705950 was in the opposite direction (i.e. the allele associated with increased risk of IPF was protective for severe COVID-19)^6^.

We aimed to further explore the shared genetic architecture and identify novel shared genetic loci between the two diseases, using new enlarged GWAS of IPF and COVID-19 risk.

## Methods

### Data

We used the largest GWAS of IPF risk which consisted of unrelated European individuals from across five studies^5^. Cases were selected from centres in the USA, UK and Spain diagnosed using American Thoracic Society and European Respiratory Society guidelines.

For COVID-19, the summary statistics from version 6 of the COVID-19 Host Genetics Initiative (HGI_v6, https://www.COVID-19hg.org/results/r6/) were used. This analysis considered four different COVID-19 phenotypes according to the severity of the disease and the controls used; A2) Very severe respiratory confirmed COVID-19 vs. population, B1) Hospitalised COVID-19 vs. not hospitalised COVID-19, B2) Hospitalised COVID-19 vs. population and, C2) COVID-19 vs. population.

### Genetic Overlap

Genome-wide genetic correlation analyses were performed using LD Score Regression. Twenty previously reported IPF genetic association signals^4,5^ were investigated for their association in the four COVID-19 GWAS, and 26 variants reaching genome-wide significance in the COVID-19 GWAS were tested for their association with IPF (proxy variants – r^2^>0.8 in European population – were investigated if the top associated variant was not included). Genetic loci showing an association with both traits (after Bonferroni correction for multiple testing) were investigated to determine whether the same causal variant was driving both the IPF and COVID-19 associations using coloc^7^. Regions with a posterior probability>80% of there being a shared causal variant (assuming up to one causal variant for each trait in the region and that variant has been measured) were deemed to have colocalised.

### Identification of putative causal genes and pleiotropic effects

We investigated whether shared genetic signals were associated with gene expression in lung tissue (GTEx_lung^8^, n=515) and whole blood (eQTLGen^9^, n=31,684). Where the variant met a false discovery rate of 5%, colocalisation analyses were performed using coloc and deemed to be linked to gene expression if the posterior probability of a shared causal variant was greater than 80%.

We performed a phenome-wide association study (PheWAS) to identify if the overlapping genetic signals had been previously reported for association with other traits (p<10^−5^) using publicly available resources (PhenoScanner_v2, GWAS Catalog and Open Targets). Colocalisation analyses were performed to determine if the same causal variant was driving both traits. For overlapping signals for traits of relevance, we additionally assessed gene expression in all GTEx tissues, as described above.

## Results

There was a significant weak positive genome-wide correlation between IPF and severe COVID-19 (A2 r^2^=0.274 p=0.0045, B1 r^2^=0.279 p=0.0093, and B2 r^2^=0.261 p=0.0005) but not with COVID-19 infection (C2 r^2^=0.066 p=0.433).

Four genetic association signals showed evidence of a shared causal variant between IPF and at least one COVID-19 phenotype (posterior probability>80%), namely loci at 7q22.1, near *MUC5B*, near *ATP11A* and near *DPP9* (**Table 1a**). The 7q22.1 locus has not previously been reported for association with COVID-19. Three additional IPF genetic signals (at 17q21.31, *DSP* and *DEPTOR*) showed an association with COVID-19 but did not colocalise, suggesting there are different causal variants between the two traits at these loci. Visual inspection of the 17q21.31 locus revealed extended linkage disequilibrium (due to the presence of a large inversion) meaning colocalisation analyses could not determine whether there were shared or distinct causal variants.

**Table 1:** Variants reaching Bonferroni-corrected significance for both IPF and COVID-19. All results are presented in terms of the allele which increases risk of IPF. Chr=chromosome. REF=reference allele. EFF=effect allele (i.e. the variant the effect estimates are in relation to. OR=odds ratio. CI=confidence interval. FEV_1_=forced expiratory volume in 1 second. FVC=forced vital capacity. HbA1c=Haemoglobin Type A1c. **a) Genome-wide analysis results for IPF and COVID-19**. The coloc column gives the posterior probability there is a shared causal variant between IPF and that COVID-19 phenotype at that genetic loci. Colocalisation analyses were only performed on signals showing a possible association with both traits after correcting for multiple testing. Signals that colocalise (i.e. posterior probability>80%) between IPF and COVID-19 are highlighted in grey. **b) Gene expression and PheWAS results**. Percentages shown for the eQTL columns show the posterior probability of colocalisation between the IPF risk signal and the gene expression eQTL signal (only genes with posterior probability>80% are presented in the table). eQTL results are presented in terms of the IPF risk allele. For the PheWAS results, phenotypes where the variant had p<10^−5^ and which colocalised with the IPF signal (posterior probability>80%) are presented. Only non-ILD and COVID-19 phenotypes were investigated for the PheWAS analysis. Proxy variants (with r^2^>0.8) were also investigated in PhenoScanner. For Open Targets only traits with genome-wide summary statistics from GWAS Catalog were investigated.

**a) Genome-wide analysis results for IPF and COVID-19**
| Chr | Position | rsid | Previously reported gene | REF | EFF | IPF |  | A2) Very severe respiratory confirmed COVID-19 vs. population |  |  | B1) Hospitalised COVID-19 vs. not hospitalised COVID-19 |  |  | B2) Hospitalised COVID-19 vs. population |  |  | C2) COVID-19 vs. population |  |  |
| --- | --- | --- | --- | --- | --- | --- | --- | --- | --- | --- | --- | --- | --- | --- | --- | --- | --- | --- | --- |
|  |  |  |  |  |  | 4,125 cases vs 20,464 controls |  | 8,779 cases vs 1,001,875 controls |  |  | 14,408 cases vs 73,191 controls |  |  | 24,274 cases vs 2,061,529 controls |  |  | 112,612 cases vs 2,474,079 controls |  |  |
|  |  |  |  |  |  | OR [95% CI] | p | OR [95% CI] | p | Coloc | OR [95% CI] | p | Coloc | OR [95% CI] | p | Coloc | OR [95% CI] | p | Coloc |
| 7 | 99630342 | rs2897075 | 7q22.1 | C | T | 1.30 [1.23, 1.37] | 1.77×10<sup>-21</sup> | 1.07 [1.04, 1.12] | 1.63×10<sup>-4</sup> | 88.2% | 1.02 [0.99, 1.05] | 0.238 | - | 1.04 [1.02, 1.06] | 5.55×10<sup>-4</sup> | 48.1% | 1.01 [1.00, 1.02] | 0.014 | - |
| 11 | 1241221 | rs35705950 | MUC5B | G | T | 5.06 [4.67, 5.47] | 9.09×10<sup>-418</sup> | 0.83 [0.77, 0.89] | 1.17×10<sup>-7</sup> | 100% | 0.89 [0.84, 0.94] | 2.20×10<sup>-5</sup> | 98.5% | 0.89 [0.86, 0.93] | 1.22×10<sup>-8</sup> | 100% | 0.99 [0.98, 1.01] | 0.448 | - |
| 13 | 113534984 | rs9577395 | ATP11A | G | C | 1.29 [1.21, 1.38] | 4.78×10<sup>-14</sup> | 0.90 [0.87, 0.94] | 4.38×10<sup>-6</sup> | 99.0% | 0.94 [0.90, 0.97] | 8.76×10<sup>-4</sup> | 52.1% | 0.94 [0.91, 0.96] | 8.67×10<sup>-7</sup> | 99.5% | 0.99 [0.98, 1.00] | 0.037 | - |
| 19 | 4717672 | rs12610495 | DPP9 | A | G | 1.28 [1.21, 1.36] | 2.56×10<sup>-16</sup> | 1.20 [1.15, 1.26] | 1.64×10<sup>-15</sup> | 97.9% | 1.08 [1.04, 1.11] | 1.73×10<sup>-5</sup> | 98.5% | 1.11 [1.09, 1.14] | 6.09×10<sup>-18</sup> | 97.9% | 1.03 [1.02, 1.04] | 5.10×10<sup>-10</sup> | 98.1% |

**b) Gene expression and PheWAS results**
| Chr | Position | rsid | Gene expression<br>(coloc posterior probability of shared causal variant) |  |  | PheWAS |  |  |
| --- | --- | --- | --- | --- | --- | --- | --- | --- |
|  |  |  | EFF | GTEx (Lung, n=515) | eQTLGen (whole blood, n=31,684) | PhenoScanner v2 | GWAS Catalog | Open Targets |
| 7 | 99630342 | rs2897075 | T | - | Decreased *ZKSCAN1* (99.4%)<br>Decreased *TRIM4* (85.4%) | Blood traits<br>Mean corpuscular haemoglobin<br>Red cell distribution width<br>Impedance of leg right | Chronic obstructive pulmonary disease | Lung function<br>FEV<sub>1</sub>/FVC<br>Peak expiratory flow<br>Blood traits<br>Mean corpuscular haemoglobin<br>Red cell distribution width<br>Red blood cell count<br>Mean corpuscular volume<br>Mean corpuscular haemoglobin concentration<br>Platelet count<br>Mean platelet volume<br>Low density lipoprotein cholesterol levels |
| 11 | 1241221 | rs35705950 | T | Increased *MUC5B* (100%) | - | - | - | - |
| 13 | 113534984 | rs9577395 | C | - | Increased *ATP11A* (99.6%) | Blood traits<br>Red cell distribution width<br>HbA1c | - | Blood traits<br>Mean corpuscular volume<br>Mean corpuscular haemoglobin<br>Red cell distribution width<br>Platelet count<br>Red blood cell count<br>Lung function<br>FEV<sub>1</sub>/FVC |
| 19 | 4717672 | rs12610495 | G | - | Decreased *DPP9* (88.3%) | - | - | Appendicular lean mass |

Three of the four shared signals colocalised with expression of the single nearest gene in blood or lung (*MUC5B, ATP11A* and *DPP9*) (**Table 1b**). The IPF and COVID-19 risk increasing alleles at the 7q22.1 signal colocalised with decreased expression of *ZKSCAN1* and *TRIM4* in blood.

The signal on chromosome 7 was associated with a number of blood traits and the signal near *ATP11A* was associated with blood traits and HbA1c (average blood glucose levels, used in diagnosing diabetes) (**Table 1b**). The IPF and HbA1c signals did not colocalise, however, as diabetes is a risk factor for COVID-19^10^, we further investigated the effects on gene expression for this signal. The allele (rs423117_T) associated with higher Hb1AC levels was associated with increased *ATP11A* expression in liver and decreased expression in cultured fibroblasts, but there was no association with *ATP11A* expression in blood.

## Discussion

Genetic association signals near *MUC5B, DPP9* and *ATP11A* have previously been reported for both COVID-19 severity and IPF risk; we show for the first time that these signals are likely due to the same underlying causal variant. In addition, we report a novel overlapping signal at 7q22.1 implicating *TRIM4* and *ZKSCAN1*.

Despite a positive genome-wide genetic correlation between IPF risk and the COVID-19 severity phenotypes (A2, B1 and B2), we show that two of the four shared signals (at *MUC5B* and *ATP11A*) have opposite directions of effect on risk for the two diseases. The allele associated with increased risk of IPF and increased *ATP11A* expression in blood (rs9577395_C) was associated with decreased risk of severe COVID-19.Our PheWAS highlighted a potential link with HbA1c and diabetes risk at this locus via *ATP11A* expression, although effects were tissue dependent.

The rs2897075_T allele at 7q22.1, associated with increased IPF and COVID-19 risk, was linked to decreased *TRIM4* and *ZKSCAN1* expression. *TRIM4* is an important regulator of virus-induced interferon induction pathways and a proteomic study identified significant adjacency between SARS-CoV-2 M protein and *TRIM4*^11^. Viral infection-induced micro-injury to the alveolar epithelium is thought to be a trigger for development of IPF^12^, suggesting the interferon-mediated innate immune response could be central to both risk of chronic lung disease and worse outcomes due to SARS-CoV-2 infection.

Loci previously implicated by IPF GWAS relating to telomere dysfunction (*TERT, TERC, RTEL1*) and mitotic spindle assembly (*KIF15, MAD1L1, SPDL1, KNL1*) were not associated with COVID-19.

The colocalisation analyses assume a single measured causal variant. Although conditional analyses found no evidence of multiple independent association signals at the regions studied, we cannot guarantee all causal variants were measured. Furthermore, we utilised whole blood and lung tissue for gene expression so we cannot rule out cell-specific effects. Further analyses in non-European populations could help identify other ancestry specific overlapping variants and increase the generalisability of the results.

In conclusion, using the largest IPF and COVID-19 GWAS to date, we show there is a positive genetic correlation between IPF and severe COVID-19 risk. However, some IPF-related pathways may have an opposite (e.g. *MUC5B* and *ATP11A* pathways) effect on severe COVID-19 risk.

## Data Availability

This study used previously published publicly available data obtained from obtained from: https://github.com/genomicsITER/PFgenetics (IPF genome-wide summary statistics), https://www.covid19hg.org/results/r6/ (COVID-19 genome-wide summary statistics), https://gtexportal.org/home/datasets (lung eQTL data) and https://www.eqtlgen.org/index.html (blood eQTL data).

## References

1. Drake TM, Docherty AB, Harrison EM, et al. Outcome of hospitalization for COVID-19 in patients with interstitial lung disease. an international multicenter study. American journal of respiratory and critical care medicine. 2020;202(12):1656–1665.

2. Pairo-Castineira E, Clohisey S, Klaric L, et al. Genetic mechanisms of critical illness in covid-19. Nature. 2021;591(7848):92–98.

3. Fingerlin TE, Murphy E, Zhang W, et al. Genome-wide association study identifies multiple susceptibility loci for pulmonary fibrosis. Nat Genet. 2013;45(6):613–620.

4. Dhindsa RS, Mattsson J, Nag A, et al. Identification of a missense variant in SPDL1 associated with idiopathic pulmonary fibrosis. Communications biology. 2021;4(1):1–8.

5. Allen RJ, Stockwell A, Oldham JM, et al. Genome-wide association study across five cohorts identifies five novel loci associated with idiopathic pulmonary fibrosis. medRxiv. 2021(12.06.21266509).

6. Fadista J, Kraven LM, Karjalainen J, et al. Shared genetic etiology between idiopathic pulmonary fibrosis and COVID-19 severity. EBioMedicine. 2021;65:103277.

7. Giambartolomei C, Vukcevic D, Schadt EE, et al. Bayesian test for colocalisation between pairs of genetic association studies using summary statistics. PLoS genetics. 2014;10(5):e1004383.

8. GTEx Consortium. Genetic effects on gene expression across human tissues. Nature. 2017;550(7675):204.

9. Võsa U, Claringbould A, Westra H, et al. Unraveling the polygenic architecture of complex traits using blood eQTL meta-analysis. BioRxiv. 2018:447367.

10. Singh AK, Gupta R, Ghosh A, Misra A. Diabetes in COVID-19: Prevalence, pathophysiology, prognosis and practical considerations. Diabetes & Metabolic Syndrome: Clinical Research & Reviews. 2020;14(4):303–310.

11. Meyers J, Ramanathan M, Shanderson R, et al. The proximal proteome of 17 SARS-CoV-2 proteins links to disrupted antiviral signaling and host translation. bioRxiv. 2021.

12. John AE, Joseph C, Jenkins G, Tatler AL. COVID-19 and pulmonary fibrosis: A potential role for lung epithelial cells and fibroblasts. Immunol Rev. 2021.

